# Longitudinal IgG antibody responses to *Plasmodium vivax* blood-stage antigens during and after acute vivax malaria in individuals living in the Brazilian Amazon

**DOI:** 10.1101/2022.08.30.22279402

**Authors:** Tenzin Tashi, Aditi Upadhye, Prasun Kundu, Chunxiang Wu, Sébastien Menant, Roberta Reis Soares, Marcelo Urbano Ferreira, Rhea J. Longley, Ivo Mueller, Quyen Q. Hoang, Wai-Hong Tham, Julian C. Rayner, Kézia KG Scopel, Josué C. Lima Junior, Tuan M. Tran

## Abstract

**Background:** To make progress towards malaria elimination, a highly effective vaccine targeting *Plasmodium vivax* is urgently needed. Evaluating the kinetics of natural antibody responses to vaccine candidate antigens after acute vivax malaria can inform the design of serological markers of exposure and vaccines.

**Methodology/Principal Findings:** The responses of IgG antibodies to 9 *P. vivax* vaccine candidate antigens were evaluated in longitudinal serum samples from Brazilian individuals collected at the time of acute vivax malaria and 30, 60, and 180 days afterwards. Antigen-specific IgG correlations, seroprevalence, and half-lives were determined for each antigen using the longitudinal data. Antibody reactivity against Pv41 and PVX_081550 strongly correlated within each of the four time points. The analysis identified robust responses in terms of magnitude and seroprevalence against Pv41 and PvGAMA at 30 and 60 days. Among the 8 *P. vivax* antigens demonstrating >50% seropositivity across all individuals, antibodies specific to PVX_081550 had the longest half-life 100 days (95% CI, 83—130 days), followed by PvRBP2b (91 days; 95% CI, 76—110 days) and Pv12 (82 days; 95% CI, 64—110 days).

**Conclusion/Significance:** This study provides an in-depth assessment of the kinetics of antibody responses to key vaccine candidate antigens in Brazilians with acute vivax malaria. Follow-up studies are needed to determine whether the longer-lived antibody responses induced by natural infection are effective in controlling blood-stage infection and mediating clinical protection.

**AUTHOR SUMMARY:** To successfully eliminate malaria, highly effective vaccines against the two major human malaria species, *Plasmodium falciparum* and *Plasmodium vivax*, will be needed. Vaccines against the blood form of malaria generate antibodies that target specific proteins on the *Plasmodium* parasite to reduce its replication within the host. Studying the antibody response after natural malaria infection can help identify blood markers of parasite exposure and also shed light on the magnitude and longevity of antibodies to vaccine candidate proteins. We performed a study to determine the frequency, magnitude, and longevity of natural antibody responses against nine *P. vivax* vaccine candidate proteins in patients with vivax malaria in Brazil. These proteins were selected based on prior studies demonstrating that antibodies against these proteins were either associated with protection against vivax malaria or have been tested as blood markers of recent infection with vivax malaria. We identify specific vivax proteins that produce more frequent and longer-lived antibody responses in this population.

## INTRODUCTION

Malaria remains a significant public health threat, with ∼241 million malaria cases globally in 2020 [1]. While *Plasmodium falciparum* is the most prevalent malaria parasite in Africa, *P. vivax* remains the predominant malaria parasite in most countries across the Asia Pacific and in the Americas [1]. Moreover, severe, life-threatening illness caused by *P. vivax* is no longer considered a rare event [2, 3]. Although the WHO recommended widespread use of the first licensed malaria vaccine, RTS,S, this vaccine acts against *P. falciparum* and is not expected to be cross-protective against *P. vivax*. Thus, to make progress towards global malaria elimination, a highly effective vaccine targeting *P. vivax* is urgently needed.

The ideal malaria blood-stage vaccine is capable of inducing a specific and durable immune response against antigens that are essential for parasite survival. Parasite-specific antibodies can play a critical role in naturally acquired protective immunity against blood-stage malaria by disrupting the interactions between parasite ligands and cognate host receptors required for RBC invasion [4-6]. Screens of IgG antibody responses against *P. vivax* antigens with potential roles in erythrocyte binding and/or invasion in Papua New Guinea children living in areas highly endemic for *P. vivax* identified or confirmed several antigens as vaccine candidates, including erythrocyte binding protein (EBP), Duffy binding protein region II (DBPII), RBP1a, RBP2b, PVX_081550, Pv12, and Pv41 [7, 8]. Antibodies to PvRBP2b have also recently been associated with protection against clinical malaria in low-transmission settings in Brazil and Thailand [9].

Studying antibody dynamics during and after naturally acquired acute infection can identify serological markers of pathogen exposure and also shed light on the magnitude and longevity of natural humoral immunity to vaccine candidate antigens, which can in turn facilitate the vaccine development process. A recent screen of antibody responses against 342 *P. vivax* proteins in longitudinal clinical cohorts identified serological exposure markers that accurately predicted recent *P. vivax* infection [10]. Characterizing the natural IgG responses to these antigens in other malaria-transmission settings would provide further information about how their natural immunogenicity may influence vaccine responses. For instance, pre-existing antibodies generated against an antigen from prior infections can shape and potentially interfere with the antibody response generated by a vaccine targeting that antigen [11, 12]. In addition, identifying antigenic targets that naturally elicit robust and durable antibody responses can help focus vaccine development on candidate antigens that would benefit from boosting during natural exposure to the pathogen [13, 14]. To this end, we conducted a study to evaluate the prevalence, magnitude, and longevity of naturally acquired IgG antibody responses against 9 *P. vivax* vaccine candidate antigens in Brazilian patients with acute vivax malaria and who were longitudinally followed for a period of up to 180 days. Antigens were chosen based on prior evidence that naturally acquired IgG antibody responses against them have been associated with protection against vivax malaria [7-9] and/or have been validated as serological markers of recent *P. vivax* infection [10].

## METHODS

### Ethics statement

The original study for the collection of human studies was approved by the Institutional Review Board (IRB) of the Federal University of Juiz de Fora, Brazil (protocol #262.875/2013) [15]. Written informed consent was obtained from all the study participants. Use of de-identified samples and subject metadata from the cohort study was approved as exempt human subjects research by the Indiana University IRB. Plasma used as malaria-naïve controls were obtained with informed consent as part of a healthy donor protocol approved by the Indiana University IRB (protocol #1601403732).

### Study site and participants

Details of the study site and participants have been described previously [16]. Briefly, between May 2013 and August 2014, 47 study participants with acute, uncomplicated *P. vivax* malaria were enrolled and longitudinally followed up to 180 days. Participants were recruited from four farming settlements in northwestern Brazil. Three settlements were in the municipalities of Acrelândia, Nova Califórnia, and Plácido de Castro [16]. A fourth settlement, Jéssica, was situated near Acrelândia. This region, located in the Amazon Basin just north of the Bolivian border, has low-level, year-round *P. vivax* transmission with no reported cases of *P. falciparum* malaria since early 2012 [17, 18]. Criteria for inclusion were children and adults aged 12 to 64 years with acute, febrile vivax malaria diagnosed first by thin smear with further single species-specific confirmation via real-time PCR targeting the *pvr47* and *pfr364* genes [16]. Participants were confirmed to be free of *Plasmodium* parasitemia by microscopy and real-time PCR at days 30, 60, and 180 after enrollment, and none of the participants had additional laboratory-confirmed malaria episodes between enrollment and their final visit. Exclusion criteria were chronic illness; history of immunodeficiency or use of immunosuppressive medications; recent malaria or use of anti-malarial medication in the 30 days prior to acute malaria presentation; and pregnancy.

### Sample collection

Venous blood samples were collected into heparinized tubes at the time of admission for acute vivax malaria (Day 0, n=44) and on post-admission days 30 (n=40), 60 (n=22), and 180 (n=21) for those participants who were able to complete follow up. At the time of enrollment, all acute vivax cases were treated with anti-malaria drugs (25 mg/kg of chloroquine over 3 days and 0.5 mg/kg/d of primaquine for 7 days) in accordance with the Ministry of Health of Brazil’s guidelines. Plasma was separated from blood cells by centrifugation within 6 h of collection, aliquoted, and stored at -80°C until use in August 2021. Blood samples from 12 malaria-naïve North American volunteers were used as non-malaria exposed controls.

### Recombinant proteins and coupling

Full-length *P. vivax* ectodomains for specific antigens (accession numbers listed in **Table S1**) were obtained from previously published plasmids deposited in Addgene [19], or, in the case of PVP01_0102300, synthesized specifically for this project. The PVX_088910 and PVX_113775 ectodomains were subcloned into a pTT3 expression plasmid so that they were preceded by mouse variable κ light chain signal peptide [20], and followed by an enzymatic biotinylating sequence and a hexa-histidine tag. The PVX_097720, PVX_110810, PVX_000995 and PVX_081550 were subcloned into a similar pTT3 expression plasmid, but in which contains a rat Cd4 tag preceding the Bio-linker and hexa-histidine tag. The constructs were expressed as described earlier [21], transfected using a 1:2.5 ratio of vector with PEI (40kDa). To produce biotinylated proteins, 1.25 μl *E. coli* BirA ligase plasmid was co-transfected with the plasmid of interest. The DNA/PEI mixture was then incubated for 8 min and transfected into HEK 293E cells. The cells were cultured in Freestyle 293 media (Invitrogen) supplemented with Geneticin (Gibco, USA), Biotin (4.8 mg/ml) and 1% fetal bovine serum (Gibco, USA) and were maintained in a shaker flask at 125 rpm, 37°C, 5% CO_2_ and 70% relative humidity. After 5 days the cell supernatant was harvested and subjected to Ni-NTA chromatography using 1 ml HisTrap column (Sigma-Aldrich) with 1 ml/min flow rate. For subsequent binding and washing, 20 mM phosphate buffer with 25 mM imidazole, pH 7.4 was used. The proteins were eluted using 20 mM phosphate buffer with 250 mM imidazole, pH 7.4. The eluants were concentrated using a 10 kDa Vivaspin concentrator (Millipore). The concentration was measured in a NanoDrop (Thermo Fisher Scientific), and purified proteins were frozen at -80ºC until further use. The PvRBP recombinant proteins were expressed with relevant tags as previously described [6, 22, 23] (**Table S1**). The sequences of all *P. vivax* recombinant proteins were based on the genome of the *P. vivax* Sal-1 strain, a monkey-adapted parasite line originally isolated from a *P. vivax* patient in El Salvador [24].

The control proteins bovine serum albumin (Sigma-Aldrich) and tetanus toxoid (Calbiochem/Millipore Sigma) were obtained from commercial vendors. Each *P. vivax* antigen or control protein was coupled to one color-coded magnetic Luminex® microsphere bead regions. The coupling procedure was followed as per manufacturer’s recommended protocol (xMAP® Antibody Coupling Kit, Luminex Corporation) using 40 pmol of each protein per 10^6^ magnetic beads, which ensured bead saturation for proteins of different molecular weights. After coupling and final wash, all protein-coupled beads were combined into a single bead mixture containing an approximately equal concentration of coupled beads for each protein (∼650 coupled beads for each protein/μl).

### Multiplex immunoassay

The multiplex bead-based immunoassay was performed per the manufacturer’s protocol with adaptation using previously published methods [25, 26]. Briefly, 96-well, flat-bottom, black microplates were pre-washed with phosphate-buffered saline (pH 7.4) containing 0.01% Tween-20, and 0.05% sodium azide (PBS-TN). Plasma was diluted 1:400 in PBS pH 7.4 containing 0.01% Tween-20 and 1% BSA (PBS-T) prior to adding to a mixture of protein-coupled beads. Samples were randomized to plates by subject with time points from the same subject maintained on the same plate. Assays were run at a final reaction volume of 100 μl per well containing diluted plasma and 1 μl of each protein-coupled bead. Plasma and beads were mixed and incubated at RT at 550 RPM for 45 mins. After washing with PBS-TN, 100 μl of phycoerythrin-labelled donkey anti-human IgG (Abcam) diluted 1:500 in PBST were added to each well. Samples were incubated for 45 min in the dark with constant shaking. The beads were re-suspended in 150 μl PBS-TN prior to acquisition and analysis using the Multiplex MAGPIX system running xPONENT 4.1 software (Luminex Corporation). For this study, the positive reactivity control consisted of pooled plasma from 20 acute vivax plasma samples randomly selected from the study participants. Plasma samples from 12 malaria-naïve North American donors were used as negative reactivity controls for determining seropositivity thresholds, defined as mean reactivity + 2 standard deviations of naïve donors. Prior to downstream analysis, for each antigen, data was normalized across different plates by multiplying fluorescent intensities by a normalization factor that was equal to the median value of all plates divided by the median value of the plate. Initially, background reactivity against rat Cd4 was subtracted from all values on a per subject basis for antigens with a rat Cd4 fusion. However, a considerable proportion of individuals exhibited background reactivity to rat Cd4 that was higher than against the vivax recombinant proteins. Therefore, we used subtracted background reactivity to BSA for all antigens.

### Statistical analyses and modeling of antibody kinetics

Analysis was performed using R version 4.0.4. Tests for significance are noted in the figure and table legends where appropriate. To more accurately model antibody decay using a simple exponential model, the maximum antibody reactivity was used as the starting point for the decay curve for each antigen and each subject. By excluding earlier time points with lower reactivity than the maximum, only the downward-sloping portion of the natural antibody response during acute malaria was utilized. Subjects without any response during acute infection or less than two data points were excluded from the regression model. For each antigen, linear mixed effects regression was performed using the lme4 [27] and lmerTest [28] packages to estimate IgG antibody half-lives, with ln(reactivity+1) as the dependent variable and time (days) and subject as fixed and random effects, respectively. Half-life (_t1/2_) was calculated as ln(0.5)/estimate. Additional analyses were also performed with the addition of age (years), sex, city, and years of residence in the malaria-endemic area as fixed effects. For years of residence in the endemic area, missing data for two subjects were imputed using the median.

The same approach was used for the re-analysis of the Longley et al. dataset [29]. However, only the model using days and subject as fixed and random effects, respectively, was performed, and analysis was limited up to 24 weeks (168 days) to maintain consistency with the current study’s maximum follow-up time of 180 days. Visualization was performed using the ggplot2 and ggpubr packages.

### Data and code availability

Data and code are available on https://github.com/TranLab/Brazil-Pv-IgG-Kinetics-2022/.

## RESULTS

### Participant characteristics

Plasma samples were obtained from Brazilian individuals of all ages who presented with acute febrile vivax malaria (**Table 1**). Individuals from Nova Califórnia and Jéssica had on average been resident in the Amazon basin for a significantly longer time than those near Acrelândia and Plácido de Castro (**Table 1**, p=0.033), suggesting different levels of prior exposure. However, there were no differences in gender proportions, age, or number of prior malaria episodes between sites. The number of individuals who completed the 30-, 60-, and 180-day follow-up visits were 37, 19, and 8, respectively.

**Table 1.** Baseline characteristics of participants by study site.

| Variable | Overall | Acrelândia | Nova<br>Califórnia | Jéssica | Plácido de<br>Castro | p |
| --- | --- | --- | --- | --- | --- | --- |
| n | 47 | 10 | 6 | 19 | 12 |  |
| Male gender (%) | 34 (72.3) | 7 (70.0) | 3 (50.0) | 15 (78.9) | 9 (75.0) | 0.576 |
| Age, years (median [range]) | 32.0<br>[12.0, 64.0] | 31.0<br>[14.0, 55.0] | 41.0<br>[29.0, 56.0] | 30.0<br>[18.0, 56.0] | 23.0<br>[12.0, 64.0] | 0.183 |
| Number of prior malaria<br>episodes (median [range])<sup>1</sup> | 3.0<br>[0.0, 30.0] | 1.0<br>[0.0, 6.0] | 3.0<br>[0.0, 15.0] | 3.0<br>[0.0, 30.0] | 2.0<br>[0.0, 10.0] | 0.211 |
| Years residence in Amazon<br>basin (median [range])<sup>1</sup> | 26.0<br>[7.0, 64.0] | 21.0<br>[8.0, 43.0] | 33.0<br>[7.0, 54.0] | 30.0<br>[18.0, 54.0] | 19.0<br>[7.0, 64.0] | 0.033 |

Numbers are n. (%) unless otherwise noted. Fisher's exact test was used for categorical variables. Kruskal-Wallis Rank Sum Test was used for continuous variables. <sup>1</sup>Self-reported at enrollment.

### Correlation of antibody responses between antigens

Pairwise correlation of IgG reactivity across all time points revealed strong and highly significant positive correlations (defined here as Spearman ρ>0.70, adjusted p-value <0.01) between Pv12, Pv41, PvGAMA, and PVX_081550 at the time of presentation (day 0; **Figure 1**). Notably, IgG reactivity against Pv41 and PVX_081550 strongly correlated across each of the four time points, suggesting that these two antigens induce similar longitudinal responses across individuals. Strong and highly significant positive correlations were also observed between PvEBP and Pv41 at day 60; between PvEBP and PvGAMA at day 180; and between Pv12 and PVX_081550 and Pv41 at day 180. Additional significant correlations are shown in **Figure 1**.

**Figure 1.**
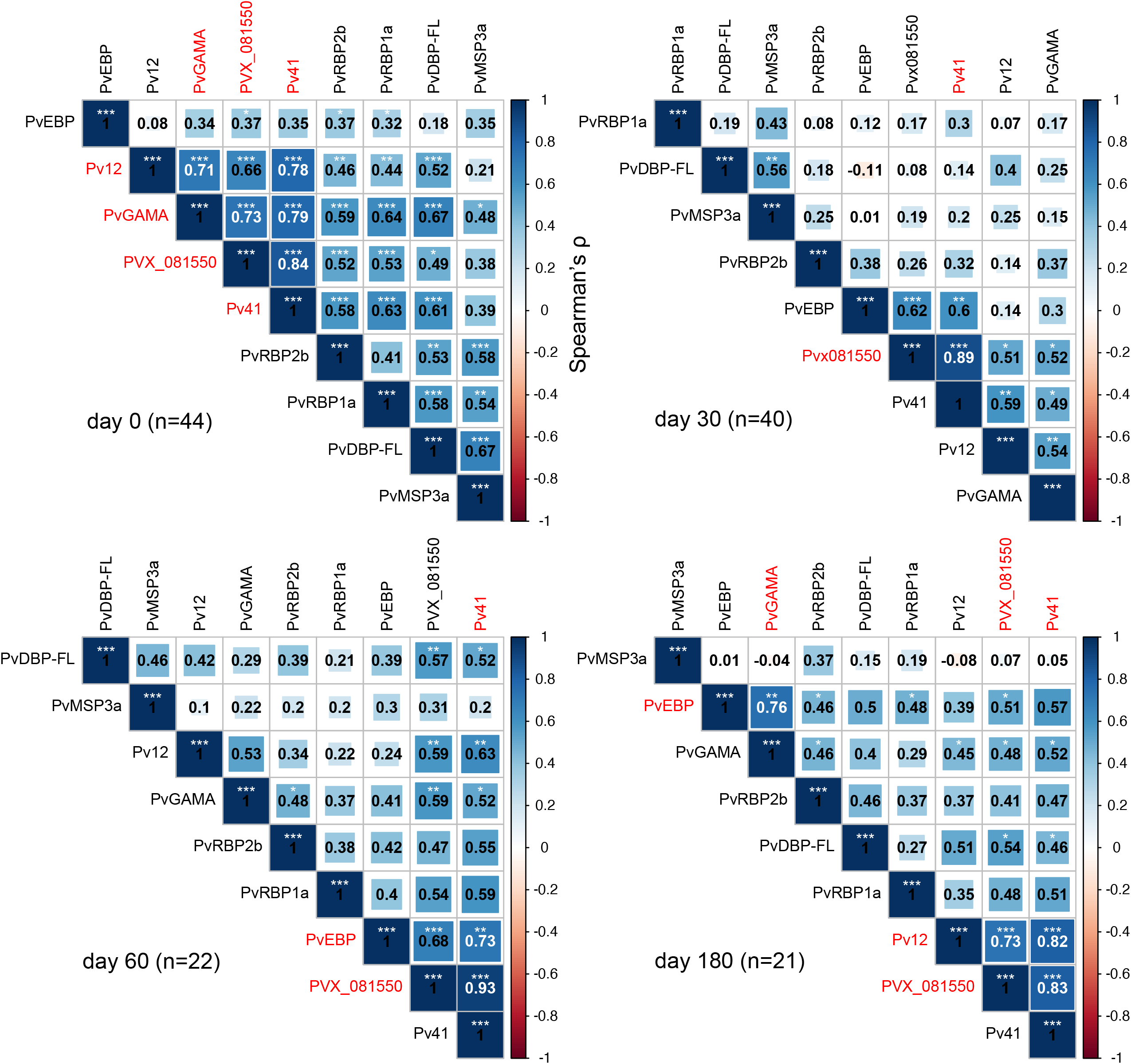
Antibody reactivity correlations between each *P. vivax* antigen by time point. Pairwise Spearman’s correlations were determined between normalized IgG reactivities for each antigen at day 0 (acute malaria presentation), 30, 60, and 180. Correlation matrices were clustered using the ward.D2 hierarchical agglomerative clustering method. Shown are Spearman’s ρ and significance adjusted for multiple testing using the Holm method. * <0.05, **<0.01, ***<0.001 Red text indicates antigen pairs that have a Spearman ρ > 0.70 and adjusted p-value<0.01, which suggests a strong positive and highly significant correlation.

### Longitudinal seropositivity rates

To assess the maintenance of antibody responses after acute vivax malaria, seropositivity rates were determined for each antigen at each time point (**Figure 2**). The majority (>50%) of acutely infected individuals became seropositive by day 30 for all antigens except for PvRBP1a. At days 30 and 60 after acute vivax malaria, 100% and 90-91% of individuals were seropositive to Pv41 and PvGAMA, respectively. At 180 days after acute vivax, >70% of individuals remained seropositive against the individual antigens Pv41, PvGAMA, and PvRBP2b, with 100% of individuals seropositive to at least one of these three antigens (**Figure 2**). By contrast, PvRBP1a was less naturally immunogenic in this population, with only 9-15% seropositivity within the first 30 days of acute infection and 0% seropositivity by 180 days (**Figure 2**).

**Figure 2.**
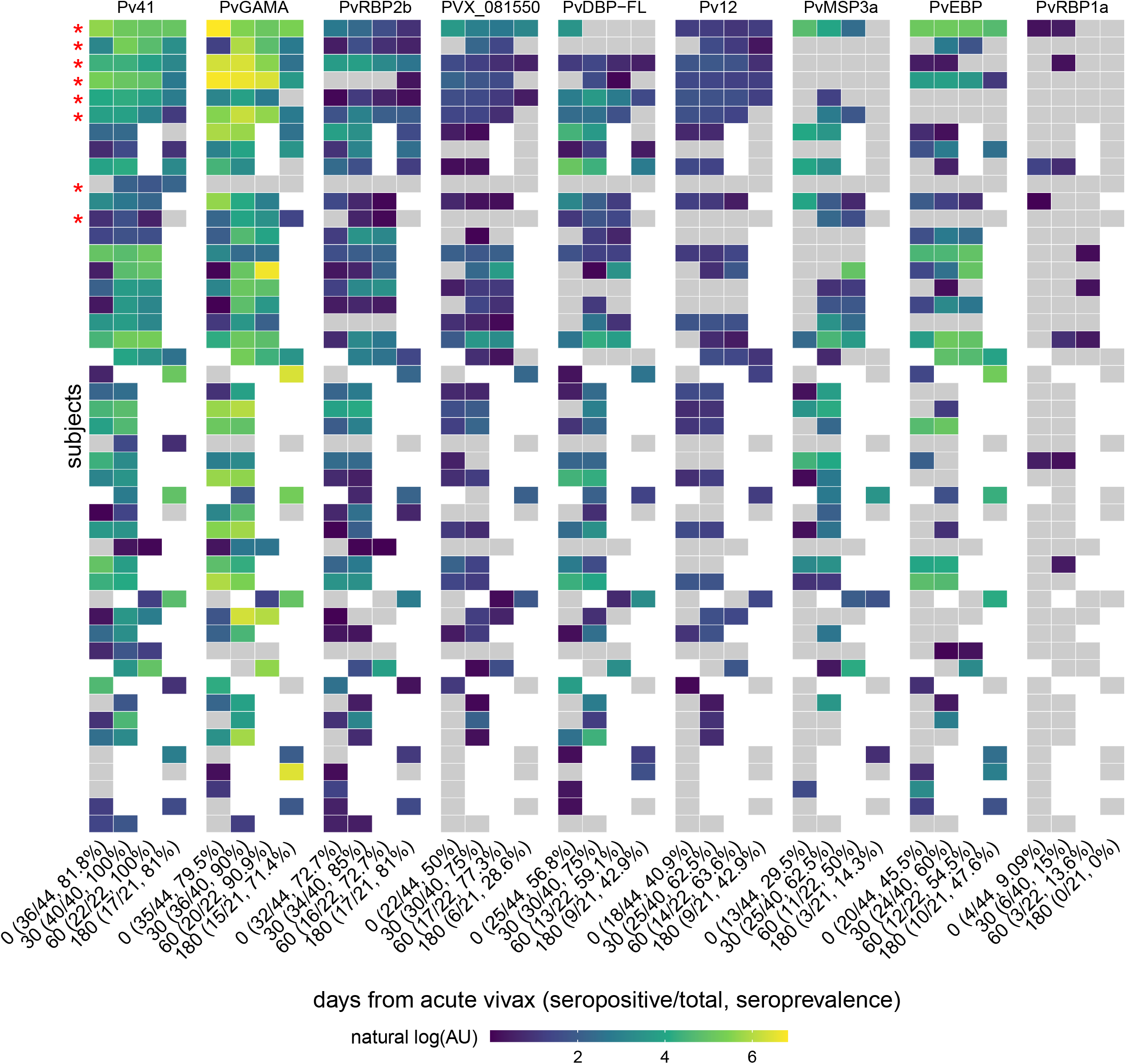
Seropositivity by antigen by time point. For each antigen, a seropositivity cut-off was set as the mean reactivity of malaria-naïve North American control samples + 2 standard deviation. Reactivities were converted to arbitrary units (AU) where by fluorescence intensity for each antigen was divided by its respective seropositivity cut-off. Antigens are ordered from left to right by seroprevalence, and columns within each antigen are ordered by seroprevalence. Rows, representing subjects, are ordered from top to bottom by overall seropositivity followed by completeness of longitudinal samples. Gray tiles represent samples that were seronegative (AU≤1). Blank tiles are missing samples. Red asterisks indicate subjects with data at all four time points.

### Kinetics of *P. vivax* antigen-specific responses post-infection

For each of the 8 *P. vivax* antigens that elicited positive responses in >50% of individuals, we estimated the decrease in specific IgG post-infection using a simple exponential decay model, based on the assumption of relatively constant antibody clearance after malaria treatment and in the absence of reinfection. To better fit this model, we used the time point having maximum antigen-specific IgG reactivity for each individual as the origin. Tetanus toxoid was used as a non-malaria-specific comparator. Antibodies against Pv12, PvRBP2b, PVX_81550, and Pv41 demonstrated slower decay rates than antibodies to PvGAMA, PvEBP, PvDBP-FL, and PvMSP3a (**Figure 3A**). The exponential model better explained time-dependent variation in antibody decay for Pv41, PvRBP2b, and PvGAMA than for the other antigens, as suggested by adjusted R^2^ values > 0.60 (**Figure 3B**). Not surprisingly, the exponential model did not explain variation in tetanus-specific IgG reactivity as well as for the *P. vivax* antigens given the variable (and unknown) time since tetanus toxoid immunization for individuals in the study.

**Figure 3.**
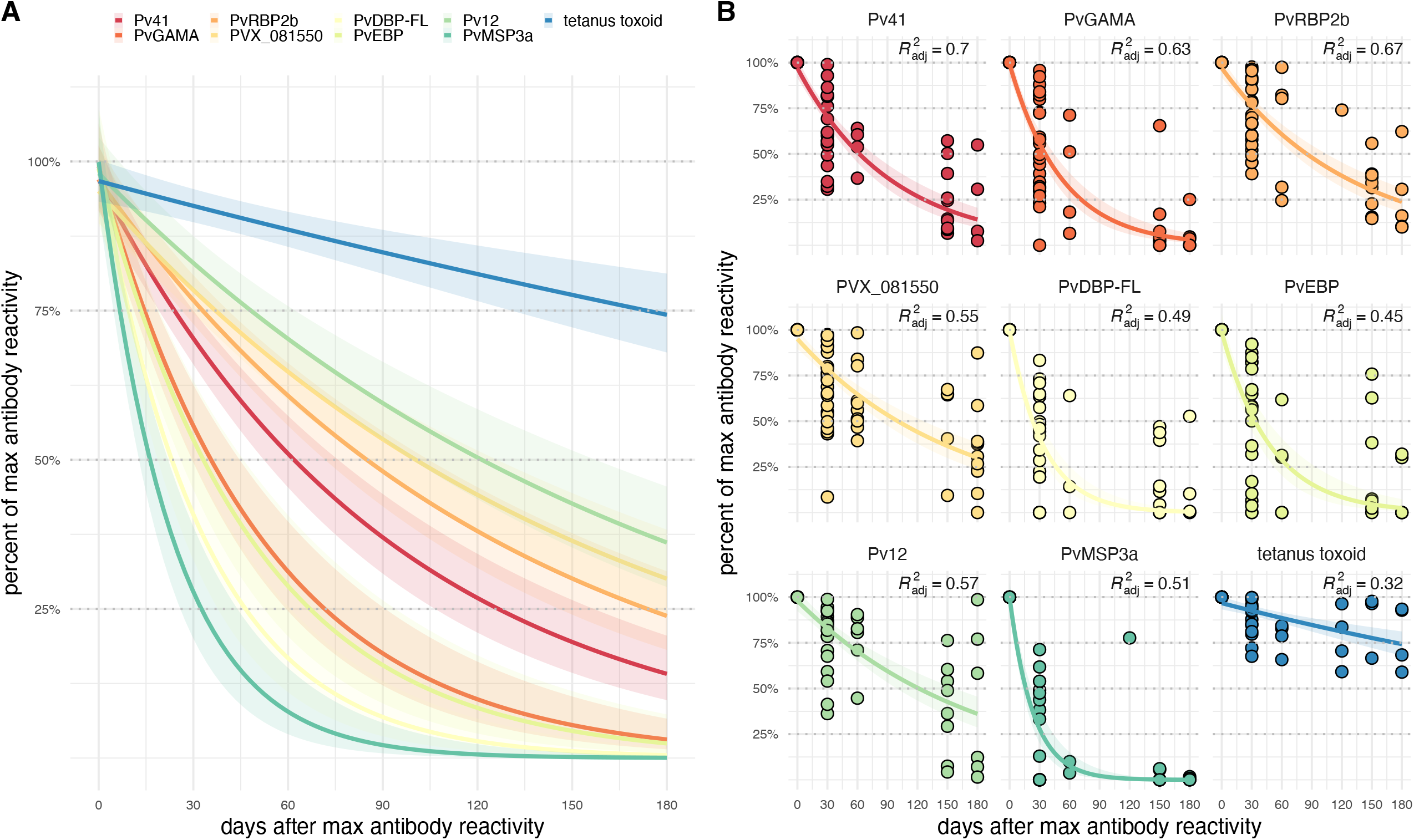
Kinetics of antigen-specific IgG responses after acute vivax malaria. **(A)** Exponential fit curves for antibody reactivities normalized as the percent of maximum antigen-specific IgG across all time points for each individual and for each antigen and plotted against days from the timepoint with maximum antibody reactivity. Bands represent 95% confidence intervals. **(B)** Same exponential fit curves in (A) with actual reactivity data. A small perturbation of 0.0001 was added to percent of maximum reactivity to avoid taking the log of 0.

### Estimation of antibody half-lives

Linear mixed-effects regression was used to estimate antigen-specific IgG half-lives. Antibody half-life for tetanus toxoid was estimated as 430 days (95% confidence interval [CI], 310—690 days; **Table 2**). Among the 8 *P. vivax* antigens evaluated, antibodies specific to PVX_081550 had the longest half-life of 100 days (95% CI, 84—140 days), followed by PvRBP2b (91 days; 95% CI, 76—110 days) and Pv12 (82 days; 95% CI, 64— 110 days). By contrast, antibodies against PvDBP-FL, PvMSP3a, and PvGAMA had significantly shorter half-lives of <45 days (**Table 2**). An adjusted analysis controlling for age, gender, prior malaria episodes, years of residence in the endemic area, and municipality was also performed with minimal change in estimates (**Table S2**). Unadjusted IgG half-life estimates were compared to longitudinal antibody kinetic data from a prior study in which plasma from Brazilians with acute vivax malaria were screened against >300 *P. vivax* proteins [29, 30]. The Longley et al. dataset was generated using a high-throughput method (AlphaScreen assay) that utilizes crude proteins and does not standardize protein concentrations between different antigens, which may explain the wider discrepancies in antibody half-life estimates for some antigens [29]. Comparisons were made using the published IgG half-lives [30], as well as IgG half-lives that were re-calculated using the same parameters as in **Table 2**. Analysis was limited to *P. vivax* antigens that were assayed in both studies and demonstrated overall seropositivity during acute vivax malaria in both studies. These parameters excluded PvRBP1a and PvEBP. Overlapping 95% confidence intervals for IgG antibody half-lives were observed between the current study and the re-analyzed Longley et al. dataset for the antigens with longer IgG half-lives (PVX_081550, PvRBP2b, Pv12, and Pv4) but not for antigens with shorter IgG half-lives (PvDBP-FL, PvGAMA, and PvMSP3a) (**Figure 4**).

**Table 2.** Antibody half-life estimates using linear mixed-effects regression of antibody reactivity.

| Antigen | estimate | standard error | statistic | $t_{1/2}$ (days) | lower 95% CI | upper 95% CI |
| --- | --- | --- | --- | --- | --- | --- |
| tetanus toxoid | -0.0016 | 0.00031 | -5.3 | 430 | 310 | 690 |
| PVX_081550 | -0.0067 | 0.00079 | -8.5 | 100 | 83 | 130 |
| PvRBP2b | -0.0076 | 0.00072 | -11.0 | 91 | 76 | 110 |
| Pv12 | -0.0085 | 0.00120 | -7.3 | 82 | 64 | 110 |
| Pv41 | -0.0110 | 0.00100 | -11.0 | 64 | 54 | 79 |
| PvEBP | -0.0140 | 0.00190 | -7.0 | 51 | 39 | 71 |
| PvDBP-FL | -0.0160 | 0.00230 | -6.9 | 44 | 34 | 63 |
| PvMSP3a | -0.0170 | 0.00220 | -7.7 | 41 | 32 | 55 |
| PvGAMA | -0.0210 | 0.00180 | -12.0 | 32 | 28 | 39 |

A linear mixed effects regression with $\ln(\text{reactivity})$ as the dependent variable and time (days) and subject as fixed and random effects, respectively. Half-life ( $t_{1/2}$ ) was calculated as $\ln(0.5)/\text{estimate}$ .

**Figure 4.**
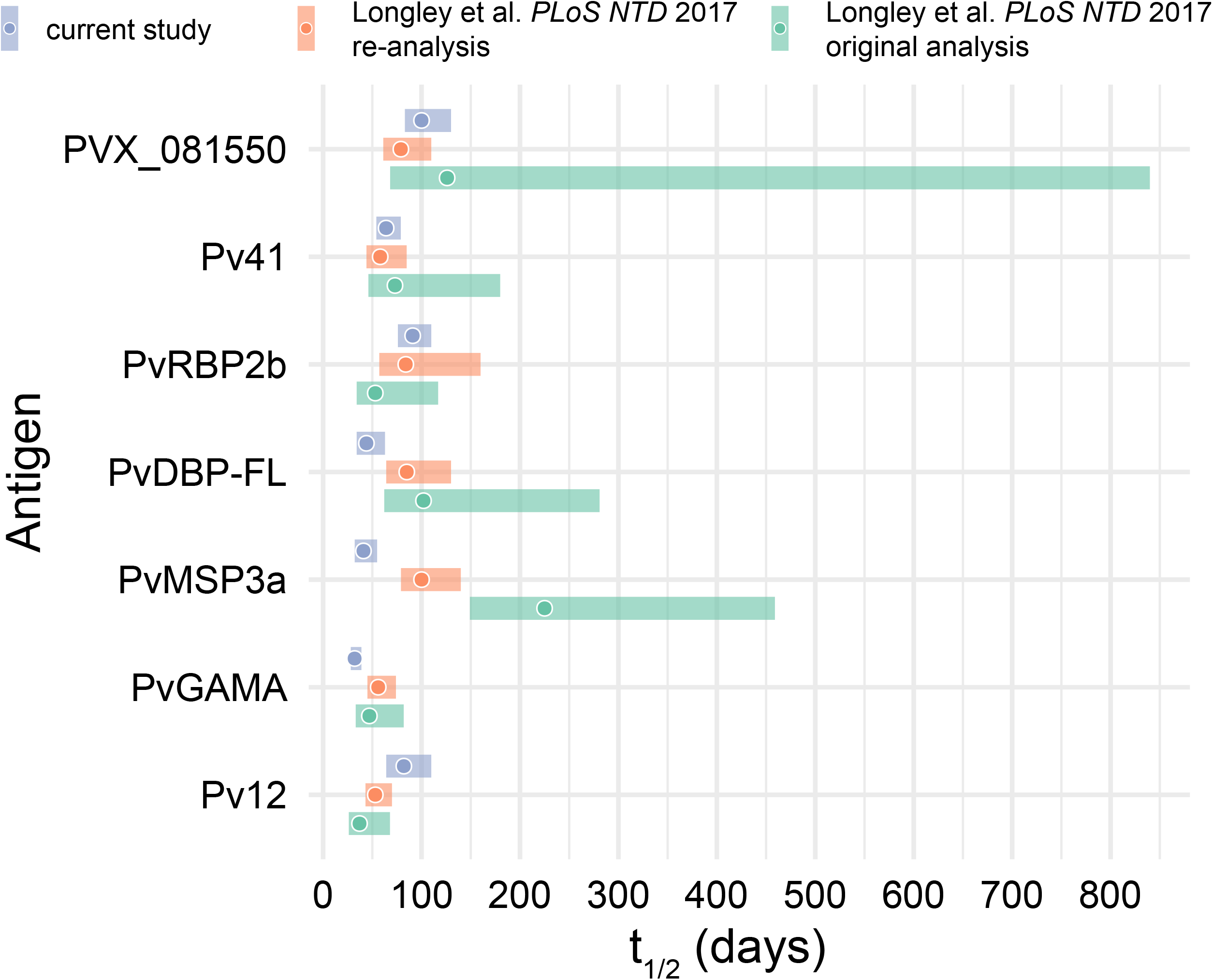
Comparison of IgG half-life estimates for *P. vivax* blood-stage vaccine candidate antigens. Reported and estimated antigen-specific IgG antibody half-lives for the Longley et al. dataset and the current study. Points are half-lives estimated from linear mixed effects models (see Methods). Bands represent 95% confidence intervals. The re-analysis of the Longley et al. dataset only utilized data to 24 weeks to more closely match the current study, whereas the original analysis of the Longley et al. dataset used all available data to 9 months.

## DISCUSSION

Assessing the dynamics of antibody responses to vaccine candidate antigens after natural infections can aid our understanding of the protective host response and facilitate rational vaccine design. In this study, the kinetics of the IgG antibody response to 9 protective antigens after acute vivax malaria were determined by following Brazilian patients up to 6 months after treatment in the absence of any known re-infections. Acute vivax malaria elicited robust IgG responses against Pv41, PvGAMA, PvRBP2b, PVX_081550, and PvDBP-FL, with at least 75% of individuals seropositive at 30 days after presentation. Such high reactivity is consistent with studies that demonstrated their robust immunogenicity in the context of natural vivax infections [19, 31]. Antibody responses of the greatest magnitude and breadth in terms of fold-change seroprevalence, respectively, were observed for Pv41 and PvGAMA. This may have implications for the degree of boosting one might expect after natural infection for potential vaccines that target either of these antigens.

Antibody responses against PVX_081550, which putatively encodes a StAR-related lipid transfer protein, demonstrated the longest half-life (100 days) of the antigens tested, consistent with the long-lived kinetics for anti-PVX_081550 antibodies observed in cohorts of Thai and Brazilian individuals with acute vivax [10, 29]. Antibodies against PvRBP2b and Pv12 also demonstrated relatively long half-lives of >80 days. The durable IgG responses to these protective antigens after acute vivax malaria may explain why clinical immunity to *P. vivax* is acquired more rapidly than to *P. falciparum* [32], as asymptomatic *P. vivax* infections can often be observed even in low-endemicity settings [33, 34]. By contrast, PvGAMA-specific IgG half-lives were relatively short-lived for both this study (32 days) and the Longley et al. data set (47 days) [29]. While there were a few similarities in antibody half-life estimates between the current study and the Longley et al. dataset, marked differences were observed for PvDBP-FL and PvMSP3a, which may be explained by the use of purified versus crude protein preparation in the current study and Longley et al., respectively. The narrower half-life confidence intervals estimated in the current study and the reanalysis of the Longley et al. dataset compared to its original analysis were likely achieved by limiting the analysis to data from the maximum reactivity up to 168 or 180 days, which removes the highly variable data points prior to the peak and near the tail of the antibody response curve.

It is important to distinguish between comparisons of antibody kinetics after acute vivax using half-life estimates and the maintenance of seropositivity. The latter is dependent on the magnitude of antigen-specific IgG reactivity over a pre-defined threshold, in this case defined by standard deviations over the average reactivity of malaria-naïve controls. With respect to maintenance of seropositivity, the combination of Pv41, PvGAMA, and PvRBP2b may be adequate indicators of more temporally remote *P. vivax* exposure, which could prove useful for sero-surveillance in communities with low transmission [10, 35]. Of these three antigens, PvRBP2b had the longest estimated half-life (91 days). Taken together, these data are consistent with the prior study which found antibodies to PVX_094255 (the same PvRBP2b_161–1454_ fragment used here) to have 75% sensitivity and specificity for classifying recent vivax exposure [10].

The high degree of correlation (Spearman ρ = 0.84-0.93) between natural IgG responses against Pv41 and PVX_081550 over all four time points during the 6-month surveillance period is intriguing. A prior study of Papua New Guinean children also found a significant albeit more moderate correlation (Spearman ρ = 0.4) between antibody responses against these two antigens [7, 8]. Given that Pv41 and PVX_081550 share only 16% amino acid identity in the reference Sal-1 sequence using the basic local alignment search tool [36], the correlative responses are less likely explained by cross-reactivity. Pv41 is a GPI-anchored protein on the merozoite surface that forms a heterodimer with Pv12 [19], which could explain why IgG responses against these antigens were highly correlated. However, PVX_081550, which has yet to be functionally characterized [8], is not known to be associated or co-localize with Pv41. Regardless of the mechanism, such tightly coordinated antibody responses during natural infection could be exploited by the inclusion of both antigens in a multi-component blood-stage vaccine, especially since IgG responses against Pv41 and PVX_081550 have been shown to be associated with protection against malaria risk when evaluated alone or in combination with IgG responses to other *P. vivax* antigens [7, 8].

Of note, the antibody half-life for tetanus toxoid, which was used as a non-malaria reactivity control in this study, was estimated to be between 310 and 690 days (0.85 and 1.89 years) in this Brazilian population, which is more comparable to the 0.98 years estimated in malaria-exposed Malian children [37] than the 11 years observed in North American adults [38]. This provides additional evidence that exposure to malaria and other tropical infections may contribute to the waning of vaccine-specific antibody responses.

### Limitations of the study

A major limitation of the study was the lack of pre-infection baseline samples, which precluded serological estimates of prior vivax blood-stage exposure and determination of true seroconversion rates. An additional limitation was the small number of subjects with full follow-up through 180 days (n=8), which potentially explains the relatively wide confidence intervals for half-life estimates for some antigens, although they may not be as wide as other comparable studies. Additionally, the first follow-up time point after an acute presentation was 30 days, which is likely at least a couple of weeks beyond the actual peak antibody response for most individuals [39]. Longevity of IgG may be due to differential subclass reactivity, which was not assessed in this study. Lastly, determining the functional significance of a long-lived, naturally acquired antibody response to these antigens, whether by neutralizing parasite-host interactions or via effector functions, was beyond the scope of this study.

### Summary

This study has determined that several recently identified *P. vivax* vaccine candidate antigens are the target of robust, long-lived natural antibody responses induced during and after acute vivax malaria. Further studies will be required to characterize the functional activity and more precise specificities of these naturally acquired, long-lived IgG antibodies and whether such antibodies are protective against clinical malaria in this community.

## Data Availability

All data produced are available online at https://github.com/TranLab/Brazil-Pv-IgG-Kinetics-2022/.

## ACKNOWLEDGEMENTS

We would like to thank the participants of the study. We are grateful to Michael Macklin and Erik Gaskin for their technical assistance. This project was funded with support from the Indiana Clinical and Translational Sciences Institute funded, in part by Grant Number UL1TR002529 from the National Institutes of Health (NIH), National Center for Advancing Translational Sciences, Clinical and Translational Sciences Award. T.M.T was supported by NIH K08AI125682. Additional support was provided by NIH R01 AI137154 (J.C.R). The content is solely the responsibility of the authors and does not necessarily represent the official views of the National Institutes of Health. W.H.T. is a Howard Hughes Medical Institute-Wellcome Trust International Research Scholar (208693/Z/17/Z) and supported by National Health and Medical Research Council of Australia (NHMRC) (GNT1160042, GNT1154937). R.J.L. and I.M. are also supported by the NHMRC (GNT1173210 to R.J.L., GNT1092789, GNT1134989, GNT1132975 and GNT1043345 to I.M.). The original field study was supported by Fundação de Amparo à Pesquisa de Minas Gerais (FAPEMIG, APQ-00769-12) and Conselho Nacional de Desenvolvimento Científico e Tecnológico (CNPq: 470315/2012-1). R.N.R.S. was recipient of CAPES doctoral fellowship. Q.Q.H. acknowledges funding from the NIH (R01GM111639, R01GM115844) and the Michael J. Fox Foundation.

## AUTHOR CONTRIBUTIONS

Conceptualization: KKGS, JCLJ, TMT

Data Curation: RRS, MUF, KKGS

Formal Analysis: TT, AU, TMT

Funding Acquisition: RJL, IM, QQH, WHT, JCR, KKGS, JCLJ, TMT

Investigation: RRS, MUF, TT, PK, CW, SM

Resources: RRS, MUF, KKGS, WHT, JCR, JCLJ

Supervision: QQH, JCR, WHT, JCLJ, TMT

Visualization: AU, TMT

Writing – Original Draft Preparation: TT, PK, SM, TMT

Writing – Review & Editing: RJL, IM, QQH, WHT, JCR, KKGS, JCLJ, TMT

