## Supplementary material for "Longitudinal IgG antibody responses to *Plasmodium vivax* blood-stage antigens during and after acute vivax malaria in individuals living in the Brazilian Amazon": Table S1

**Table S1.** Recombinant proteins used in the study.

| **PlasmoDB accession #** | **Protein** | **Description** | **Plasmid source (addgene)** | **MW** | **His tag**  **(4.2 kDa)** | **rat CD4 tag**  **(20.4 kDa)** | **estimated total MW (kDa)** | **References** |
| --- | --- | --- | --- | --- | --- | --- | --- | --- |
| PVX_097720 | MSP3a (full length) | Merozoite Surface Protein 3a | #68509 | 90.6 | Yes | Yes | ~95 | [21] |
| PVX_088910 | PvGAMA  (full length) | *Plasmodium vivax*  GPI-anchored micronemal antigen | #68522 | 80.8 | Yes | No | ~85 | [20, 21] |
| PVX_113775 | Pv12  (full length) | *Plasmodium vivax* 12 | #68516 | 38.6 | Yes | No | ~43 | [20, 21] |
| PVX_110810 | DBP  (full length) | Duffy binding protein ectodomain | #68528 | 117.1 | Yes | Yes | ~142 | [21] |
| PVX_000995 | Pv41  (full length) | *Plasmodium vivax* 41 | #68519 | 44.1 | Yes | Yes | ~68 | [21] |
| PVP01_0102300 | EBP (DBP 2) | Erythrocyte Binding Protein/Duffy Binding Protein 2 | Gene synthesized in Twist Biosciences | 95.1 | Yes | No | ~120 | this study and  [21] |
| PVX_081550 | StAR-related lipid transfer protein, putative  (full length) | StAR-related lipid transfer protein putative | #68532 | 56.9 | Yes | Yes | ~81 | [21] |
| PVX_094255 | PvRBP2b | *Plasmodium vivax* reticulocyte binding protein 2b | - | 152.8 | No | No | ~153 | [22, 23] |
| PVX_098585 | PvRBP1a | *Plasmodium vivax* reticulocyte binding protein 1a | - | 117.9 | No | No | ~118 | [23] |
| - | Cd4 | rat Cd4 domain 3 and 4 tag | - | 20.3 | No | No | ~24.5 | Rayner Lab |
